# Prediction of Hospital Outpatient Attendance in UK Hospitals: A Retrospective Study Applying Machine Learning to Routinely Collected Data for Patients of All Ages

**DOI:** 10.1101/2022.01.24.22269733

**Authors:** Jonathan Holdship, Harpreet Dhanoa, Adrian Hopper, Claire J. Steves, Mark Butler, Ingrid Wolfe, Katie Tucker, Carolyn Cooper, Jeremy Yates

## Abstract

**Objectives:** Patient non-attendance at outpatient appointments is a major concern for healthcare providers. Non-attendances increase waiting lists, reduce access to care and may be detrimental not for the patient who did not attend. We aim to produce a model which can accurately predict which appointments will be attended.

**Setting:** A teaching hospital in London, UK combining secondary and tertiary care.

**Participants:** A set of 9.6 million outpatient appointments between April 2015 and September 2019 including all ages and specialities.

**Primary and secondary outcome measures:** Area under the receiver operating characteristic curve (AU-ROC) for prediction of outpatient appointment non-attendances.

**Results:** The model uses 27 predictors to achieve an AUROC score of 0.768 (95% CI: 0.767-0.769) and accuracy of 89.2% (95% CI: 89.16%-89.24%) on test data. We find that the waiting period between booking and the appointment, the patient’s past attendance behaviour, and the levels of deprivation in their local area are important factors in predicting future attendance.

**Conclusion:** Our model successfully predicts patient attendance at outpatient appointments. Its performance on both patients who did not appear in the training data and appointments from a different time period which covers the Covid-19 pandemic indicate it generalized well across both face to face and virtual appointments and could be used to target resources and intervention towards those patients who are likely to miss an appointment. Moreover, it highlights the impact of deprivation on patient access to healthcare

**Strengths and Limitation of this Study:**

- We make use of a large dataset which enables us to use complex machine learning algorithms.
- We validate the model on two large, distinct datasets giving high confidence in our model performance.
- An unknown amount of patient data is missing due to a nearby hospital which shares patients with the study setting.

## 1 Introduction

Outpatient services have an increasingly important role in clinical care by secondary and tertiary health-care providers. Non-attendance at booked clinic appointments is common - averaging 8% in the English NHS [1] - and may disproportionately impact disadvantaged groups or patients living with multimorbidity or frailty. This Did Not Attend (DNA) rate remains high although there is variation in England between NHS hospital trusts and by specialty within NHS hospital trusts [2].

The use of machine learning is growing rapidly in healthcare. Some applications are specific to a small group of patients or a specific technique but may not answer large scale problems where patient centred factors including those linked to health inequalities may be important. In this study we took routine data available at a hospital and national deprivation data to develop a predictive tool to identify those patients at risk of non-attendance at a booked outpatient clinic appointment.

Many factors associated with increased risk of non-attendance are also associated with increased risk of poor health outcomes so that those patients most likely to miss appointments are likely to be those most at risk of harm from disruption of their care [3]. Clinic non-attendance also causes costs associated with wasted clinic time and underutilised staff, with an average cost per COVID-19 missed appointment of £150 in the UK or almost £1 billion annually [1]. Beyond the financial costs, there are opportunity costs for other patients waiting for a clinic appointment which is particularly important when there is high demand.

Untargeted interventions to prevent non-attendance are common [4]; NHS patients routinely receive email and SMS reminders about upcoming appointments. However, given the relatively small proportion of patients who DNA, manual interventions to improve the DNA rate will be wasted on the majority of patients. Since interventions such as phone calls are only slightly more effective than SMS reminders [5], it is unclear whether untargeted, manual interventions would be cost effective. Moreover, strategies such as predictive overbooking where more appointments are scheduled than can be seen are effective but require a way to estimate how likely a patient is not to attend [6].

Therefore it is useful to identify patients who have a high risk of non-attendance. There have been attempts to address this problem using machine learning models. One group at University College Hospital [7] analysed 22,000 MRI appointments at two hospitals, training gradient boosting machines to predict DNAs. They obtained an AUROC score of 0.852. Another, used a neural network to predict appointment attendance for a wider range of appointment types and patients at the Royal Berkshire NHS Foundation Trust. They obtain an AUROC score of 0.71 [8]. Both of these studies indicate there is value in machine learning approaches to this problem as their models perform significantly better than a random approach.

In this work, we apply a similar approach to train machine learning models to predict whether a patient will attend their outpatient appointment. However, we consider a much larger patient group by using every outpatient appointment scheduled at the Guy’s and St Thomas NHS Foundation Trust (GSTT) between April 2015 and September 2019 for all ages and specialities. Guy’s and St Thomas NHS Foundation Trust (GSTT) is a large multisite teaching hospital in central London with a comprehensive range of secondary and tertiary care specialties. The local population for the hospital includes areas with high levels of deprivation and the mean outpatient DNA rate is higher than NHS average (9.8% in 2017 [9]).

The aim of this study is to develop and validate a predictive model which predicts whether an individual patient will attend a scheduled outpatient appointment. We aim to produce a general purpose model for a large tertiary healthcare provider that predicts non-attendance for all ages and specialties rather than specific clinics or patient groups.

## 2 Methods

A series of classification models were trained to classify appointments as a non-attendance or attendance event using data available up to the day before the appointment. The best performing algorithm was then taken forward to develop a final predictive model.

### 2.1 Data Pre-processing

The main dataset was a record of 9.4 million individual outpatient appointments for all specialities and for patients of all ages between April 2015 and September 2019 who were referred to GSTT. We pseudonymize the data by assigning each patient a unique ID and removing other patient ID numbers and names. The demographics of these patients are given in Table 1.

**Table 1:** Demographic breakdown of patients in the training and test data. We use the UK’s office of national statistics ethnic categories.

| Characteristic | Demographic group | Training Proportion | Test Proportion |
| --- | --- | --- | --- |
| Age | Under 18 | 9.6% | 9.4% |
|  | 18-40 | 30.9% | 30.9% |
|  | 41-65 | 37.3% | 37.3% |
|  | Over 65 | 22.1% | 22.4% |
| Sex | Female | 55.5% | 55.4% |
|  | Male | 44.5% | 44.6% |
| Ethnicity | Asian | 5.2% | 5.3% |
|  | Black | 17.4% | 17.3% |
|  | Mixed | 3.3% | 3.3% |
|  | Not Stated | 6.3% | 6.3% |
|  | Other | 3.6% | 3.5% |
|  | White | 64.1% | 64.3% |

The starting date was chosen to coincide with a change in data collection at GSTT so that all data was collected in the same way and thus the sample size is all possible records conforming to this constraint. We exclude appointments that were cancelled with notice by either the patient or GSTT. For each appointment, the dataset contained patient demographic details as well as details of the appointment. One weakness of our data is that an unknown portion of each patient’s medical history is missing due to the fact many patients also visit a nearby hospital which does not share data with GSTT.

This dataset was randomly split by patient in the ratio 80:20 to provide a training and test set respectively. We then randomly shuffled the training data set and kept only one appointment per patient in order to ensure the training data were independent. The test set was used only to produce the performance statistics presented in Section 3 and was not altered or de-duplicated in order to evaluate how the algorithm would work in practice. The distribution of important predictors in the training and test sets are presented in Tables 2 and 3. The DNA rate was *∼*10% in both sets.

**Table 2:** Comparison of binary predictor values between training and test data set as well as the target variable “patient did not attend” which indicates the appointment was not attended.

| Predictor | Training |  | Test |  | Average SHAP |
| --- | --- | --- | --- | --- | --- |
|  | False | True | False | True |  |
| New appointment | 0.68 | 0.32 | 0.67 | 0.33 | 0.08 |
| Patient is British | 0.63 | 0.37 | 0.63 | 0.37 | 0.08 |
| Appointment is non-consultant led | 0.60 | 0.40 | 0.60 | 0.40 | 0.06 |
| Appointment by self-referral | 0.91 | 0.09 | 0.91 | 0.09 | 0.06 |
| Appointment by GP referral (EBS) | 0.88 | 0.12 | 0.87 | 0.13 | 0.06 |
| Patient is female | 0.51 | 0.49 | 0.52 | 0.48 | 0.05 |
| Physiotherapy appointment | 0.90 | 0.10 | 0.90 | 0.10 | 0.04 |
| Appointment at St Thomas Hospital | 0.60 | 0.40 | 0.60 | 0.40 | 0.04 |
| Medical Specialties | 0.88 | 0.12 | 0.88 | 0.12 | 0.03 |
| Appt with Maternity Services | 0.95 | 0.05 | 0.95 | 0.05 | 0.02 |
| Urology appointment | 0.97 | 0.03 | 0.97 | 0.03 | 0.02 |
| Appointment with haemophilia clinic | 0.97 | 0.03 | 0.97 | 0.03 | 0.01 |
| Dental Services appointment | 0.90 | 0.10 | 0.90 | 0.10 | 0.01 |
| Renal appointment | 0.98 | 0.02 | 0.98 | 0.02 | 0.01 |
| Gastrointestinal Medicine and Surgery appointment | 0.95 | 0.05 | 0.95 | 0.05 | 0.01 |
| Appointment with paralysis clinic | 0.99 | 0.01 | 0.99 | 0.01 | 0.00 |
| **Patient did not attend** | 0.9 | 0.10 | 0.9 | 0.10 | - |

**Table 3:**
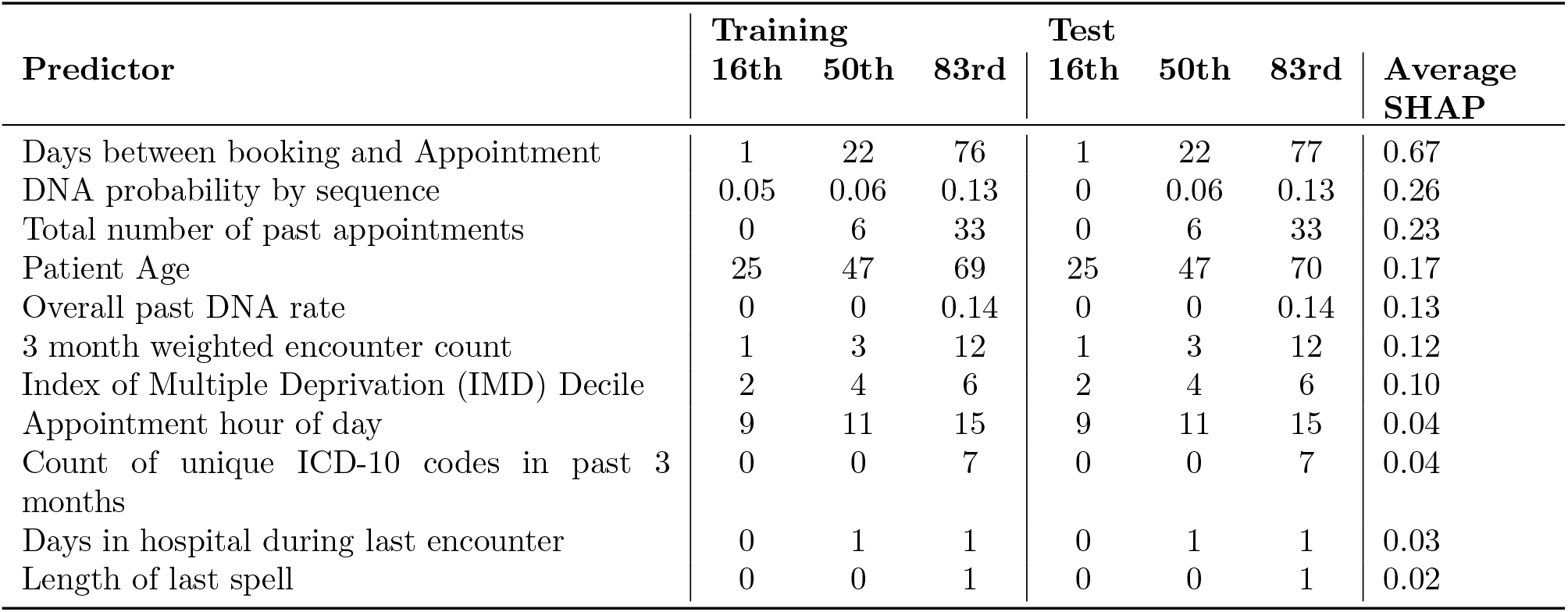
Comparison of continuous predictor values between training and test data set. We give the 16th, 50th (median) and 87th percentiles of the variable in each set. The SHAP value, indicating predictor importance is also given.

| Predictor | Training |  |  | Test |  |  | Average SHAP |
| --- | --- | --- | --- | --- | --- | --- | --- |
|  | 16th | 50th | 83rd | 16th | 50th | 83rd |  |
| Days between booking and Appointment | 1 | 22 | 76 | 1 | 22 | 77 | 0.67 |
| DNA probability by sequence | 0.05 | 0.06 | 0.13 | 0 | 0.06 | 0.13 | 0.26 |
| Total number of past appointments | 0 | 6 | 33 | 0 | 6 | 33 | 0.23 |
| Patient Age | 25 | 47 | 69 | 25 | 47 | 70 | 0.17 |
| Overall past DNA rate | 0 | 0 | 0.14 | 0 | 0 | 0.14 | 0.13 |
| 3 month weighted encounter count | 1 | 3 | 12 | 1 | 3 | 12 | 0.12 |
| Index of Multiple Deprivation (IMD) Decile | 2 | 4 | 6 | 2 | 4 | 6 | 0.10 |
| Appointment hour of day | 9 | 11 | 15 | 9 | 11 | 15 | 0.04 |
| Count of unique ICD-10 codes in past 3 months | 0 | 0 | 7 | 0 | 0 | 7 | 0.04 |
| Days in hospital during last encounter | 0 | 1 | 1 | 0 | 1 | 1 | 0.03 |
| Length of last spell | 0 | 0 | 1 | 0 | 0 | 1 | 0.02 |

A secondary data set covering all outpatient appointments from the year 2020 at GSTT was also obtained to act as a temporally distinct test set. This period covers a large part of the recent Covid-19 epidemic during which the outpatient DNA rate was 9% and many outpatient appointments were seen in virtual clinics. Thus, if the model still performs well on this data, we can be more certain of its generalisability.

### 2.2 Feature Extraction

Many variables such as patient age and the time between booking and the appointment were immediately available. Other variables required minimal processing such as the binary encoding of categorical variables including clinical specialty and patient sex. In this section, we briefly describe how more complex information was extracted from our records.

We leveraged the patient’s attendance history using the method of Goffman et al. [10]. We encoded a patient’s past appointments as a sequence of 0s and 1s where a 0 is a non-attendance and a 1 is an attended appointment. Since sequences longer than eight can be unique in our data, we only encoded up to eight past appointments. For any sequence, we can calculate the fraction of appointments in our data set where a patient had the same sequence and then did not attend. Goffman et al. found that in their data set only 20% of people with a sequence of 0000011111 (where 1s represent the most recent appointments) missed their next appointment despite having a 50% DNA rate across those eight previous appointments. Thus, the sequence provides much more insight than the overall DNA rate.

Patient postcodes were mapped to a Lower Layer Super Output Area (LSOA) which is an area of on average 650 households in order to get local deprivation information. The English Indices of deprivation rank every LSOA in seven domains: income, employment, healthcare, education, crime, and housing as well as a combined rank. The 2015 English indices of deprivation [11] were used in this work by using the patient’s LSOA to obtain their local rankings. Where a patient did not have a postcode, we used the medians from our population for their deprivation rankings.

Finally, hospital spell data for patients who had had inpatient or day case treatment over the same time period was used to augment what was known about each patient. For each appointment, the ICD-10 diagnosis codes applied to the patient over the year prior to the appointment were listed. From this, the Global Frailty Score [12] and a modified Elixhauser score [13] were calculated. Scores were assigned to all patients despite the fact that the Global Frailty Score was only validated for its original purpose on over 75s and the Elixhauser was designed for adults. Despite this, they are scores that can be calculated for any patient based on their ICD-10 code history and their potential utility as predictors in this model is independent of their validation in other contexts. Including the score for all patients is also the simplest way to include the information for adults in a way that works for models which cannot handle missing data.

### 2.3 Model Optimization and Training

In the first step, models from the Scikit-learn python library [14] as well as the XGBoost classifier [15] were trained to classify appointments using the processed data sets. They were each trained on 75% of the training data and evaluated on the remaining 25%. The AUROC score [16] on each data set was used to evaluate the models. Since not all models can handle missing data, we fill missing values in the predictors with zeroes where missing data should indicate a non-event. For example, a missing value in the length of the patient’s last spell is assumed to indicate no previous spells and so is zero filled. For other variables where a missing value certainly exists, such as the patient age, the median value of the training data is used to filling missing values.

The models and their AUROC scores are given in Table 4. Without optimization, the random forest and XGBoost classifiers outperformed other models. We therefore chose XGBoost for further optimization, favouring the fact that XGBoost can use unbalanced data to learn appropriate prior probabilities for each class whilst still learning to discriminate between classes. This is important as the data is highly unbalanced and we want the model to use the fact that 90% of appointments are attended whilst still being able to correctly identify DNA events.

**Table 4:** AUROC score obtained on validation data for four models which were trialled

| Model | Training AUROC | Validation AUROC |
| --- | --- | --- |
| K-Nearest Neighbours | 0.88 | 0.58 |
| Logistic Regression | 0.63 | 0.63 |
| Random Forest | 0.77 | 0.76 |
| XGBoost Classifier | 0.77 | 0.77 |

The first optimizing step was feature selection, we use the common approach of forward feature selection [17]. In this process, the model was first trained with all predictors to find the single predictor that produced the best single variable model. We then try all remaining predictors to find the variable that when included in a model with the first, most increases the AUROC. We do this repeatedly, adding the predictor that most improves the validation AUROC until the AUROC score becomes constant. This exhaustive approach reduces the number of predictors, removing variables that add no additional information when other predictors are already in the model.

We then use a grid search method to select the optimal values for the model hyperparameters including the learning rate, the maximum depth of the decision trees and the sample sizes used to train each tree. Those optimal values were selected by maximizing the AUROC score. To conduct the search, a 3-fold cross validation was performed using the entire training set.

## 3 Results

### 3.1 The Optimized Model

The grid search of hyperparameter space resulted in a negligible increase in the model’s AUROC score. Nevertheless, our grid resulted in an optimal learning rate (eta) of 0.01. We also limited the complexity of the model with a maximum tree depth (max depth) of 9 and min child weight of 8 resulted in marginal improvements. Finally, parameters which add randomness, making the model robust to noise were subsample with a value of 0.9 and colsample bytree of 0.7 which reflect the size of the subsample of the training data and the fraction of predictors used to train each tree respectively. Ultimately, however, the improvement in the AUROC score by choosing these parameters was of order 0.01.

### 3.2 Predictors

The fewer predictors included in the model, the more likely it is to be replicable in other settings. Of the 260 co-variates in our data set, 27 were required to obtain the same AUROC score on the training data as the full set. We assess the relative importance of predictors using SHAP values [18], an established measure of importance in machine learning models which assign importance to a predictor’s value in determining the outcome of a specific prediction using Shapley values from game theory. We have calculated the SHAP value of each predictor for every appointment in the test data. We combine these by taking the average magnitude of the SHAP values of each predictor as a measure of overall importance and give these in Tables 2 and 3. We also show the individual values in Figure 1 with colours to indicate the value of the predictor. From Figure 1, we can see how a larger (blue) or smaller (red) than average value of each predictor affects a prediction. For example, we can see short wait times between booking and an appointment have large, negative SHAP values meaning they reduce the likelihood of non-attendance but long wait times increase the likelihood.

**Figure 1:**
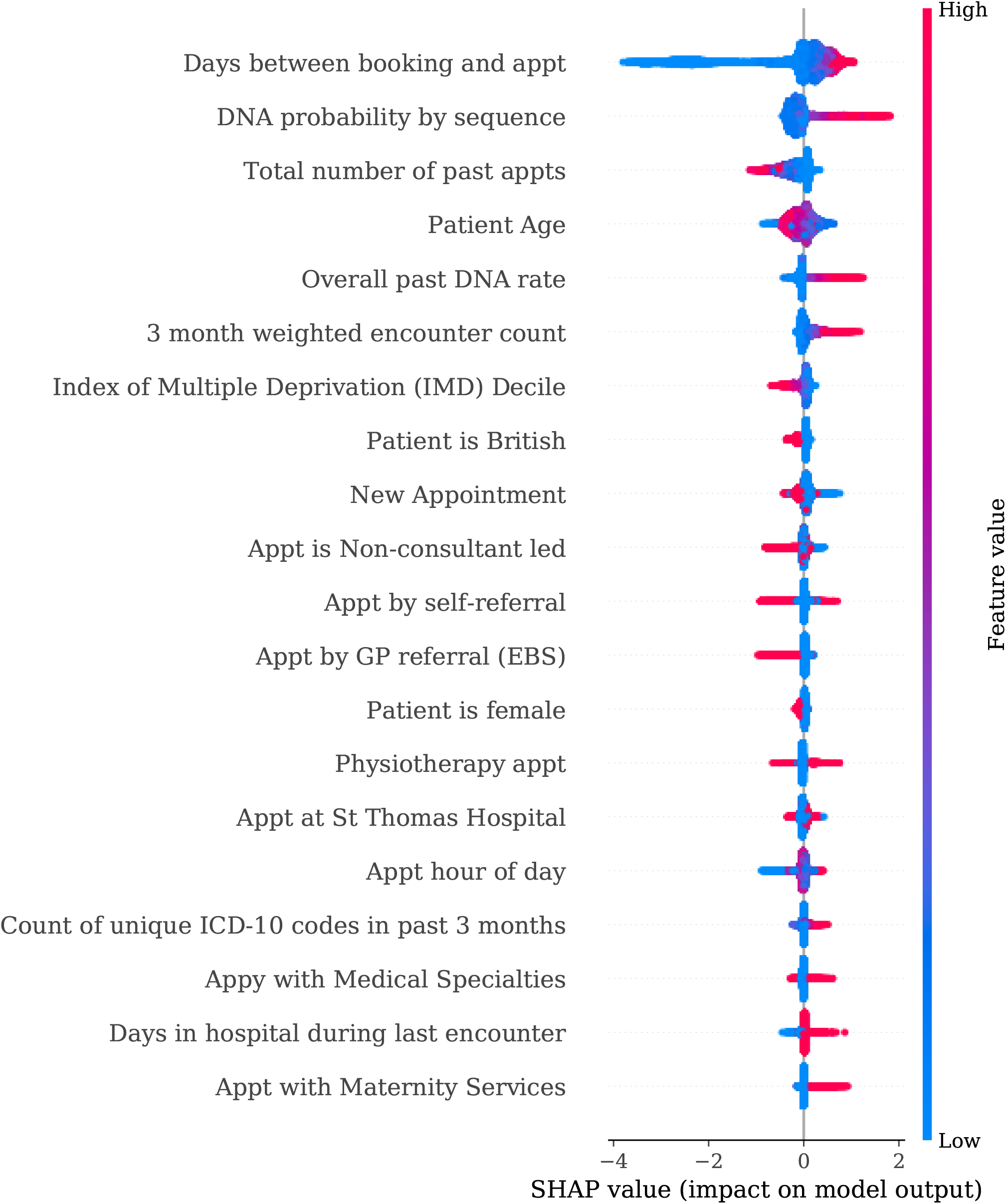
SHAP values for a subsample of the test data. Blue points represent an above average value of a predictor and red indicate a below average. The SHAP value indicates the size of the effect the predictor had on a specific prediction and whether it contribute positively or negatively to the probability of DNA. Figure 2 AUROC score as a function of the number of predictors included in the model.

In Figure 2, we show how the AUROC score obtained on the validation data improves as a function of the number of predictors included. Most interestingly, we find just six predictors can produce a model almost as accurate as the final model, with an AUROC score on the validation data of 0.75 compared to 0.76. These top six predictors include the patient’s age, total past DNA rate and the probability obtained from their attendance history sequence as well as measures of their hospital utilization over the past 3 months and the wait time between their appointment and when they booked it.

**Figure 2:**
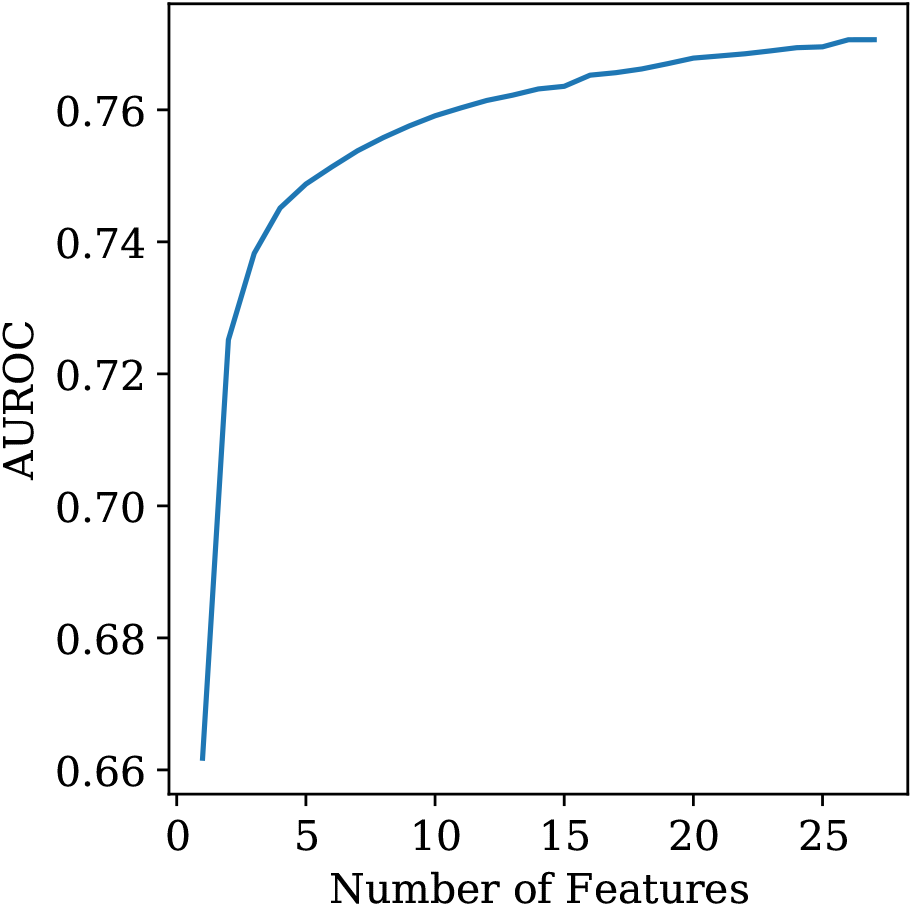
AUROC score as a function of the number of predictors included in the model.

**Figure 3:**
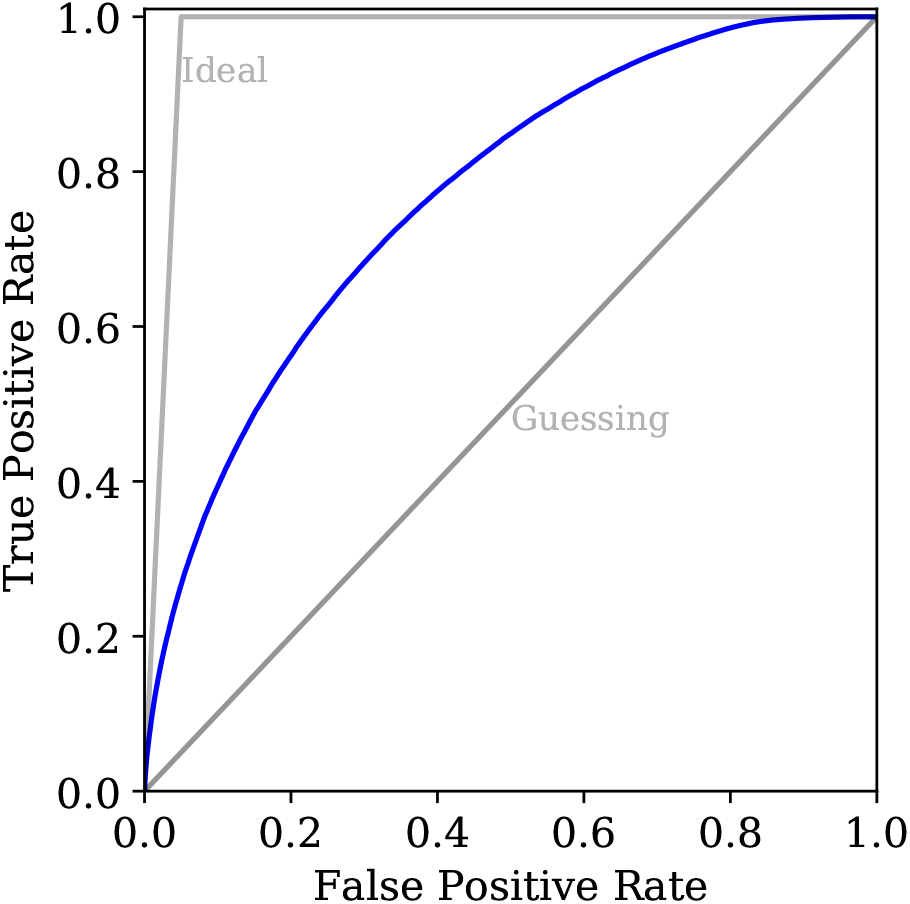
Receiver-operating characteristic curve, showing the fraction of DNA events captured as a function of the false positive rate. The grey diagonal line represents the performance of a model that makes a random guess and the blue lines indicate the false positive rate (1%) and true positive rate(10%) obtained by choosing a threshold probability of 0.8.

### 3.3 Model Performance

The final model was first tested on the 20% of data that was withheld from the training process. We estimate confidence intervals on our performance statistics using bootstrapping, calculating the metrics on 100 test sets sampled from the original set with replacement. The metrics and confidence intervals are given in Table 5. Our optimized XGBoost classifier has an accuracy of 89.2% (95% CI: 89.16 – 89.24%) which is similar to the class balance. However, this is a result of XGBoost correctly considering the prior probability of a DNA rather than simply being the spuriously high accuracy classification models can achieve by overpredicting the dominant class. We can verifying this by examining the ROC curve in Figure 5 which demonstrates the model is performing much better than a random guess. In fact, taking the area under the ROC curve, we get an AUROC score of 0.767 (95% CI: 0.766-768) indicating the model has truly learned to separate DNAs from attendances.

**Table 5:** Model performance metrics on the test data with confidence intervals from bootstrapping

| Metric | Average | 95% CI |
| --- | --- | --- |
| AUROC | 0.768 | 0.767-0.769 |
| Accuracy | 0.8920 | 0.8925-0.8924 |
| Sensitivity | 0.089 | 0.088-0.090 |
| Specificity | 0.991 | 0.9908-0.9911 |
| Positive Predictive Value | 0.550 | 0.546- 0.556 |
| Negative Predictive Value | 0.8982 | 0.8977-0.8986 |

**Figure 5:**
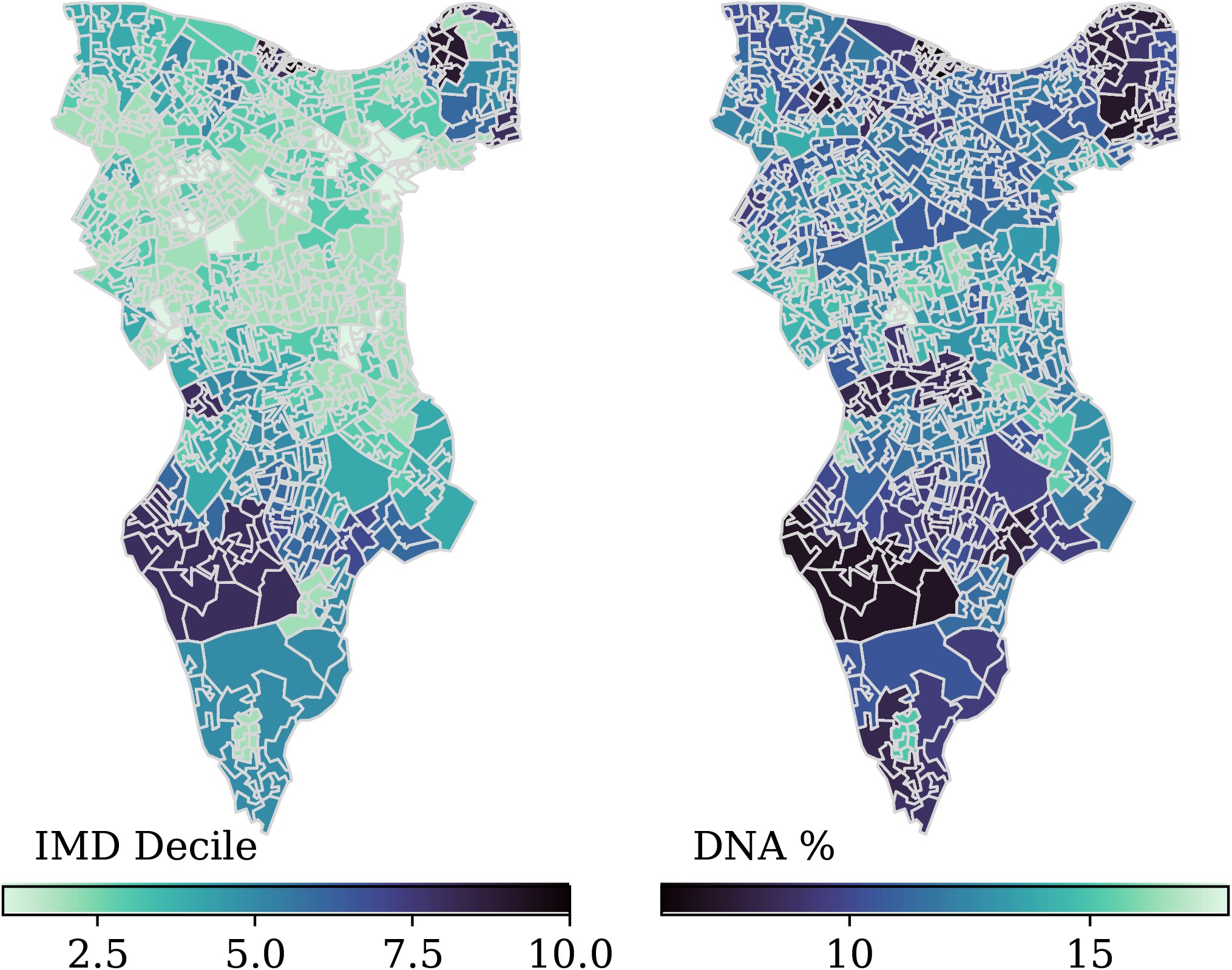
The London Borough of Southwark, with each LSOA coloured by the IMD decile (left) or DNA rate (right) of that area. Lighter colours show more deprived areas. The color scale is reversed for the DNA rate to better show the negative correlation between the two.

The AUROC compares well with the predictive model of Nelson et al. [7] which has an AUROC score of 0.85 when we consider that their model was limited to MRI appointments only and thus would have much less variance in their data set compared to our analysis. In fact, another recent work by Dashtban Li [8] uses a deep learning approach to this problem for a set of outpatient appointments more comparable to this work and obtain an AUROC score of 0.71.

The model was further tested on a set of patient appointments that were completely separated in time from the training data. Whilst the test and training data were taken from the year 2015-2019, this second test set included all outpatient appointments at GSTT in 2020. On this test set, the model achieved an AUROC score of 0.75 and so it can be concluded the model is transferable to other time periods.

Figure 4 is a normalized confusion matrix which shows that the model predicts attendance in almost every case, resulting in 10% of appointments being non-attendances that the model misses. However, of the 1.8% of appointments the model predicts are non-attendances, over half are in fact missed appointments. Thus, by training with imbalanced data, we have sacrificed sensitivity for precision that is high relative to the class balance.

**Figure 4:**
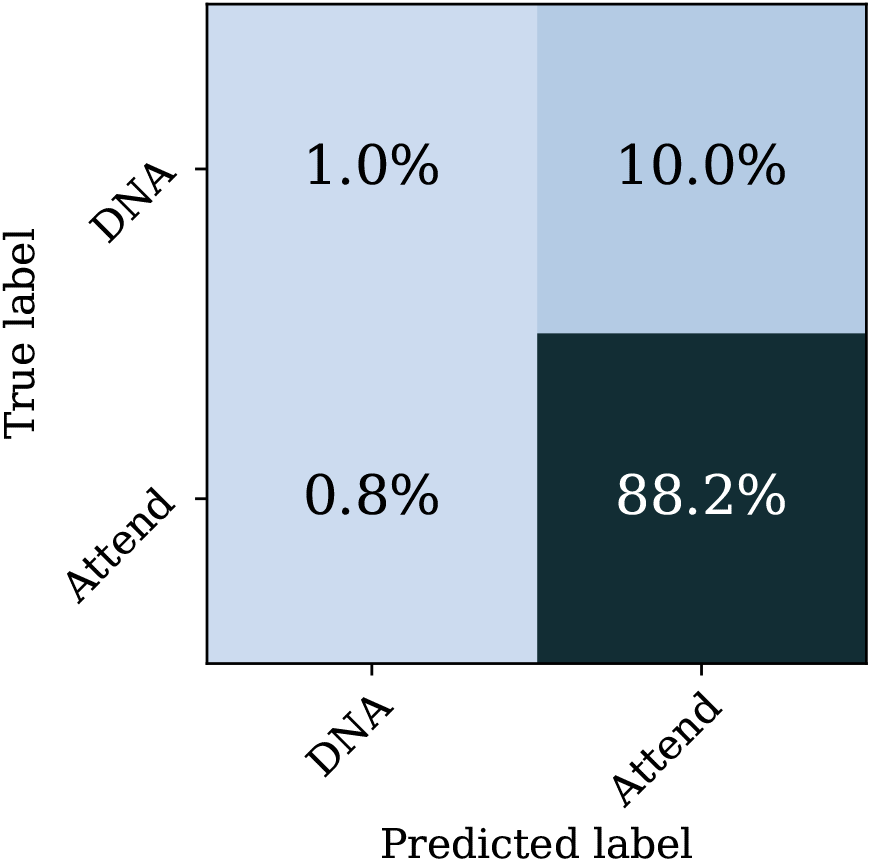
Normalized confusion matrix which shows the proportion of appointments in the test data which had a given combination of predicted and actual attendance value.

## 4 Discussion

### 4.1 Risk Stratification

The model predicts the probability of an appointment being a non-attendance which is converted to a binary prediction using a threshold. By changing that threshold, we can limit false positives at the cost of obtaining fewer true positives by demanding higher probabilities or vice versa. However, given that the model already considers the prior probability of attendance, the true positive rate can be extremely low for high thresholds. Perhaps more usefully, patients can be ranked by their probability of non-attendance. Comparing the predicted probabilities of the model to the fraction of correct predictions in the test data indicated that the model is well calibrated. i.e. we found an appointment with a predicted DNA probability of 0.90 will be missed in 90% of cases. This means that the probabilities can be used to prioritize interventions with resources first directed to those most likely to miss their appointment before moving on to contact those who are more likely to attend. The fact the model’s AUROC score is much greater than 0.5 indicates that following a strategy of first intervening in high probability cases will represent a large improvement in efficiency over randomly contacting patients.

### 4.2 Improving the Model

It is likely that some of the misclassified appointments could be correctly predicted with additional information. For example, in one early iteration of the model, the CityMapper API was queried to obtain the travel time from each postcode in London to St Thomas’ hospital for an arbitrary off-peak time to give a measure of the difficulty a patient would have in reaching their appointment. Whilst this improved the AUROC score of the model by 5%, it was dropped to limit the predictors to those that may be easily collected by tertiary care providers and not every city is covered by CityMapper. Nevertheless, it does indicate that a more specific measure of the travel time to the location of an outpatient appointment could improve the model.

Another potential area of improvement is the encoding of the patient’s diagnostic history. Measures such as the Elixhauser score use the patient’s ICD-10 coding history to determine their general health. ICD-10 codes are subject to large error rates [19] and coding is typically done with a view to generating financial data rather than predictive variables. An alternative system such as SNOMED [20] in which clinical staff directly produce diagnostic data with an aim towards predictive analytics may produce more accurate models.

### 4.3 Broader Model Applicability

This model shows a reasonable accuracy of non-attendance prediction across different time periods and across both virtual and face-to-face appointments. This allows outpatient clinics to target inventions in additional to routine care based on the patient’s predicted non-attendance risk. Each of the predictors used in the model are routinely collected in NHS hospital trusts and thus the model can be applied in other NHS hospitals in England. There is no patient information included in the random forests so they can be shared without concerns for patient privacy.

However, when moving to a different population, a more successful approach is likely to result from retraining the same algorithm using historical data from the hospital that intends to use it rather than replicating this model which results from training on GSTT data. This is due to the weighting given to each predictor. Predictors like DNA rate, ICD-10 coding rates and patient demographics vary greatly between trusts. Therefore, if the informative part of a predictor is whether a patient deviates from the average, a decision that has learned thresholds from GSTT will fail at a trust with a much different distribution.

Further, it is not possible to assume the accuracy or completeness with which each trust records any given predictor is similar. For example, the accuracy of ICD-10 coding varies greatly between trusts [21]. It’s therefore possible a model will learn to give less weight to a less accurate predictor if trained on the data from another provider.

### 4.4 The Effects of Deprivation

One of the most striking aspects of the model is the importance given to measures of deprivation. This indicates that it is the most deprived patients whose health is affected by non-attendance and that, by targeting patients for intervention, some of the effect of broader inequality on patient health could be mitigated.

Our population is an interesting one for studying the effects of deprivation; 42% of GSTT patients are from areas in the lowest 3 IMD deciles but many patients are also from the highest deciles. Figure 5 shows the London borough of Southwark which is one of the two boroughs from which the majority of GSTT patients are drawn. Each LSOA is plotted on a colour scale showing the IMD decile of that area and is compared to the DNA rate of that LSAO. The IMD ranking is ordered such that the lower deciles are the more deprived areas. These heavily overlap with the areas with the highest DNA rate. In fact, the DNA rate and IMD ranking of an LSAO have a Spearman correlation coefficient of -0.73 highlighting the need for more investigation into the link between deprivation and outpatient attendance.

## 5 Conclusion

In order to predict whether patients would attend future outpatient appointments, an XGBoost classifier model was trained on historical data from Guy’s and St Thomas NHS Foundation Trust. The input data to the model included basic appointment details, summaries of the patient medical history, their past attendance record and information about the area in which they live. The model achieved a 73% accuracy and AUROC of 0.76 in adults and children.

The model has considerable predictive power, producing predictions that are much better than random. Moreover, it performed equally well on two different test sets. The first being a set of appointments from the same time period as the training data but using a set of patients who did not appear in that training data. The second test set included all appointments from the 2019/2020 financial year which is a year after the most recent appointments in the training data and had the considerable difference of including appointments scheduled during the Covid-19 pandemic, which included a much higher proportion of ‘virtual’ outpatient appointments. Despite the model accuracy, the false positive rate is a large issue due to the small fraction of patients who actually do not attend. To counter this, the model’s output probabilities can be used rather than the binary prediction. These probabilities are well calibrated such that X% of appointments with a DNA probability of X% are missed. It is therefore expected that this model could be used to prioritize patients for intervention based on their probability of non-attendance.

We calculate SHAP values that allows us to assess which factors have the greatest effect on whether a patient will attend, finding to be clinical scheduling factors the most important. This model highlights the overlap between high LSOA deprivation and DNA rate. Any intervention should factor in the relationship between deprivation and outpatient attendance as this effect could reinforce inequalities of health outcomes which could be reinforced further by digital inequality.

Future work will focus on implementing the algorithm in clinical settings to determine the best use of the information the model provides. It is also likely the model could be improved by including auxiliary data that is not part of the routinely collected data at GSTT. The ultimate goal is to use this predictive insight into likelihood of non-attendance to improve patient management and increase healthcare efficiency. Given the importance of socio-geographic factors, at least for this London hospital, this could help reduce inequalities in healthcare. Implementation trials, for example using targeted engagement approaches for highest risk groups identified, are needed to test the best methods to integrate such prediction algorithms into local routine healthcare.

## Data Availability

The data used in this work is not available as, even in its anonymised form, it is the personal data of GSTT patients.

## Acknowledgments

JH and HD were supported by STFC DiRAC innovation fellowships. The authors also thank CityMapper for the use of their API to test whether travel times affected the model.

## Author Contributions

JH and HD contributed to the conceptualization of the study, data curation and analysis and the writing of the manuscript. AH contributed to the conceptualization of the study, interpretation of the data and critical review of the manuscript. CJS contributed to the conceptualization of the the study and interpretation of the data. MB contributed interpretation of the data and critical review of the manuscript. IW contributed interpretation of the data. KT contributed through project administration. CC contributed through project administration. JY contributed to the study methodology.

